# Gastrointestinal Dysmotility, Autonomic Function and Small Intestinal Bacterial Overgrowth Among People with Well-Controlled HIV

**DOI:** 10.1101/2024.09.25.24314370

**Authors:** Jessica Robinson-Papp, Mitali Mehta, Bridget R. Mueller, Niyati Neupane, Zhan Zhao, Gabriela Cedillo, Kaitlyn Coyle, Maya Campbell, Mary Catherine George, Emma KT Benn, Gina Lee, Jack Semler

## Abstract

**Introduction:** Gastrointestinal dysfunction, including microbiome changes and increased translocation across a compromised gastrointestinal barrier plays a role in the chronic inflammation experienced by people with HIV (PWH). It is unknown whether autonomic neuropathy (AN) may contribute to these mechanisms by altering gastrointestinal motility.

**Methods:** This is a cross-sectional study of 100 PWH and 89 controls. All participants underwent assessment of gastrointestinal transit times using a wireless motility capsule (WMC). All PWH and a subset of controls also underwent: a standardized battery of autonomic function tests summarized as the Modified Composite Autonomic Severity Score (MCASS) and its adrenergic, cardiovagal and sudomotor sub-scores, breath testing for small intestinal bacterial overgrowth (SIBO), and the Patient Assessment of Upper Gastrointestinal Disorders Symptoms (PAGI-SYM) and Composite Autonomic Symptom Score 31 (COMPASS-31) questionnaires.

**Results:** PWH displayed shorter gastric emptying times (GET) and longer small bowel and colonic transit times (SBTT, CTT) compared to controls. Among PWH, GET was associated with PAGI-SYM score. The MCASS and its sudomotor sub-score (reflecting peripheral sympathetic function) were associated with SBTT but not GET or CTT. PWH with prolonged SBTT (>6h) were more likely to have SIBO.

**Conclusion:** Gastrointestinal motility is altered in PWH. This study provides preliminary evidence that changes in autonomic function may influence SBTT in PWH and that prolonged SBTT may contribute to the development of SIBO. Future studies are needed to more fully elucidate the pathophysiologic links between HIV-associated AN, altered gastrointestinal motility, the gastrointestinal microbiome, chronic inflammation, and resulting morbidity and mortality among PWH.

## Introduction

The gastrointestinal (GI) tract plays a crucial role in HIV disease progression. Beginning in early HIV infection, there is significant depletion of CD4+ T cells from the gut mucosa which persists in chronic infection, despite effective combined antiretroviral therapy (CART).^1^ The paucity of mucosal CD4+ T cells leads to immunologic and also physical barrier disruption given the importance of CD4+ T cells in maintaining mucosal junctions. In addition, enterocytes may be lost through numerous HIV-related mechanisms,^2^ and changes in the GI microbiome may occur.^3,4^ Taken together, these changes are thought to lead to increased translocation of microbial products from the GI lumen into the systemic circulation where they serve as a source of chronic antigenic stimulus which in turn contributes to the development of medical comorbidities.^5^

Despite this broad interest in the GI pathophysiology of people with HIV (PWH), GI dysmotility has not been studied in the CART-era; prior studies included patients examined in the 1990s who were untreated or receiving zidovudine monotherapy.^6-11^ This is a potentially important omission, given that autonomic neuropathy (AN) is present in up to 60% of people with HIV,^12^ and gastrointestinal dysmotility is a common end organ effect of AN,^13^ which might contribute to dysbiosis by allowing for stasis of luminal contents.^14^ Indeed our own prior work demonstrated an association between vagal dysfunction (VD, one component of HIV-AN) and small intestinal bacterial overgrowth (SIBO) in people with HIV.^15,16^

In general, GI dysmotility might occur in AN because, although there is intrinsic rhythmicity in GI smooth muscle which is guided by pacemaker cells (interstitial cells of Cajal), normal peristalsis requires neural control. The most direct neural control is provided by the enteric nervous system (ENS), a distinct division of the autonomic nervous system (ANS) that resides within the gut and is semiautonomous. It is not possible to directly measure ENS function in humans. However, pre-clinical models demonstrate that the HIV protein Tat can alter the function of these neurons and also that HIV-related inflammation in the gut wall can damage them, leading to a gut specific HIV-AN.^17^ The ENS is also closely connected to the systemic ANS which is extrinsic to the gut. The parasympathetic/vagal branch of the extrinsic ANS tends to promote GI motility, whilst the sympathetic branch is inhibitory.^18^ Therefore dysfunction of the extrinsic ANS, as in HIV-AN, would also be expected to impact GI motility, especially in the proximal portions of the gut, such as the stomach and proximal small intestine, which are more richly innervated by the extrinsic ANS.^18^

The current study had three main goals. First, we sought to use a wireless motility capsule (WMC) that measures regional transit times throughout the entire gut to determine the prevalence of GI dysmotility among people with well-controlled HIV on combined antiretroviral therapy (CART). Second, we sought to understand whether GI motility was linked to autonomic function, and if so whether this was attributable to parasympathetic/vagal dysfunction, sympathetic dysfunction, or both. Finally, given our prior observations of high rates of SIBO among PWH, our third goal was to establish whether SIBO among PWH was potentially attributable to prolonged small bowel transit time (SBTT).

## Methods

### Overview of study design

This is a cross-sectional observational study. The original study design called for the recruitment of PWH and HIV-negative controls. However, production of the WMC was suspended in 2023 and all remaining WMCs had expiration dates in or before May 2024. Thus, the decision was made to focus on recruitment of PWH and to supplement the control group with historical control data from participants who underwent WMC testing as healthy controls in two prior studies performed by co-author JS.^19,20^ There were important demographic differences between the participants in these studies and ours. Both historic studies had fairly even recruitment of men and women, whereas our study had a male majority. Also participants in the first historic study^19^ were younger than our participants. Therefore, to more closely match our study demographics (i.e., older and more men) we used only data from male participants from the first study and all the data from the 65+ study. The historical controls did not undergo the other procedures (e.g. autonomic and breath testing) described herein.

### Participants and recruitment

All procedures were performed according to a protocol approved by the Program for the Protection of Human Subjects’ Institutional Review Board (IRB) at the Icahn School of Medicine at Mount Sinai (ISMMS). All participants provided written informed consent. All study visits were conducted at ISMMS. Participants were recruited primarily from a primary care clinic network within Mount Sinai which provides primary care to approximately 10,000 people with HIV. Potentially eligible participants were identified by pre-screening clinic providers’ schedules and then seeking permission from providers prior to contacting potential participants. Included participants were adults with well-controlled HIV (documented HIV-1 viral load <100 copies/ml within 3 months of the study visit) on stable CART for at least 3 months. Additional criteria were designed to exclude participants with a clear cause for autonomic dysfunction other than HIV (e.g., diabetes or Parkinson’s disease), those with a known history of significant GI disease, and those taking medications with significant autonomic or GI effects (e.g. opioids, sympathomimetics, prokinetics, anti-diarrheals, antibiotics). Patients for whom the required testing would present undue risk or be uninformative were also excluded (e.g., autonomic testing requires that heart rate is under sinus control and that the patient is able to stand). Criteria for the prospectively enrolled HIV-negative controls were essentially the same except they were required to have evidence of a negative HIV-test within the previous six months.

### Testing procedures

The main procedures were: WMC testing, breath testing for SIBO, and autonomic function tests (AFTs). In addition, blood, saliva and stool samples were collected and stored for future analysis.

The WMC (Medtronic) is swallowed and then passes through the entire GI tract collecting pressure, temperature, and pH data and ultimately passing during a bowel movement typically within 5 days.^21,22^ Prior to WMC ingestion, participants are instructed to hold all proton pump inhibitors and H_2_ receptor blockers for 7-10 days and fast starting at midnight the night prior. Participants consume a low fat standardized snack just prior to swallowing the WMC with 50mL of water. Participants are observed for 30 minutes and then remain fasting for 6 hours after WMC ingestion. Data are analyzed using MotiliGI software to establish three regional transit times: gastric emptying time (GET), small bowel transit time (SBTT), colonic transit time (CTT).

Autonomic function tests (AFTs) are a standard battery of non-invasive tests (WRMed), which require about 90 minutes and include: sudomotor testing (QSweat), heart rate response to deep breathing, Valsalva maneuver, and tilt.^23^ Prior to testing, participants are instructed to refrain from nicotine or caffeine consumption for at least 4 hours and avoid any medications that cause dry eyes or mouth for 24 hours. The QSweat is performed by iontophoresis of acetylcholine into the skin at four standard locations. The evoked sweat is collected and measured by an automated system. A non-invasive, continuous blood pressure (BP) monitor is then attached to the participant’s finger, and a 3-lead surface electrocardiogram and respiratory monitor are attached to the chest. Heart rate (HR), BP, and respirations are recorded continuously during: 8 slow deep breaths paced with a visual cue; forceful exhalation to a pressure of 40 mmHg for 15s (Valsalva maneuver, VM); head-up tilt for 10m. These procedures are used to calculate the Modified Composite Autonomic Severity Score (MCASS) which reflects overall autonomic function and is the sum of three sub-scores (zero is normal for each).^24^ The cardiovagal (i.e., parasympathetic) sub-score is based on changes in HR during deep breathing and VM, the adrenergic (i.e., cardiovascular sympathetic) sub-score is based on changes in BP during VM and tilt, and the sudomotor (i.e., peripheral, non-cardiovascular sympathetic) sub-score is based on the QSweat.

For the breath testing participants are instructed to consume a special diet on the day and fast overnight. Participants begin by exhaling into a standardized apparatus which collects a breath sample. They then ingest a standardized glucose solution, and repeat breath samples are then collected at 20-minute intervals for 180 minutes. The samples are analyzed in a CLIA-certified lab (Aerodiagnostics, LLC) using gas chromatography (e.g. QuinTron Micro Analyzer, QuinTron Instrument Company, Inc.). The results are expressed as maximum increase in hydrogen and methane in parts per million (ppm) and are used to assign a diagnosis of SIBO (absent vs. present) based on consensus criteria.^25^

### Medical history and patient-reported outcome measures

Medical history was obtained via chart review combined with participant interview. We calculated two measures of medical comorbidity: the Veterans Aging Cohort Study (VACS) index (v1.0 for consistency with our prior studies^26^) and the Charlson Comorbidity Index (modified to exclude HIV/AIDS).^27-29^ Participants’ medication regimens were used to calculate an anticholinergic burden (ACB) score^30^ because medications with anticholinergic effects may impact ANS function. GI symptoms were assessed using the Patient Assessment of Upper Gastrointestinal Disorders Symptoms Questionnaire (PAGI*-*SYM)^31^ which includes 20 symptoms rated on a 5-point Likert scale from “none” to “very severe” which generate a total and six domain scores: heartburn/regurgitation, fullness/early satiety, nausea/vomiting, bloating, upper abdominal pain, and lower abdominal pain.^31^ Autonomic symptoms were quantified using the abbreviated version of the Composite Autonomic Symptom Score (COMPASS-31) which includes 31 questions summarized as a total and six domain scores: orthostatic intolerance, vasomotor, secretomotor, GI, bladder, pupillomotor.^32^

### Statistical analysis

Descriptive statistics, including percents and medians with interquartile range were calculated as appropriate. Most continuous variables (including GET, SBTT, CTT, MCASS and PAGI-SYM and COMPASS-31 scores) were non-normal in distribution and so nonparametric tests were used. Median GET, SBTT and CTT were compared between PWH and control groups using the Mann-Whitney U test. Correlations between symptom scores, MCASS and transit times were assessed using Spearman’s rank correlation. Linear regression models with log-transformed outcome variables were used to assess whether between group differences or correlations persisted after adjustment for relevant co-variates. Association between prolonged SBTT (defined as >6h) and SIBO was assessed using a Chi-square test.

## Results

### Participant demographics, medical characteristics and autonomic function

Participant demographics, autonomic, GI and symptom variables are summarized in Table 1. The sample of PWH (N=100) had a median age of about 55 years and was about three-quarters male. According to inclusion criteria, they all had treated and well controlled HIV (VL<100 copies/ml); the mean CD4+ T-cell count was in the normal range. Most had longstanding HIV, with a median self-reported disease duration of 21 years. Regarding burden of co-morbid medical illness, the median VACS index was 18 (IQR 10-33) and the median Charlson was 1.0 (IQR 0-1); consistent with values found in large representative cohorts of PWH in North America.^27-29^ Regarding medications, anticholinergic burden was low overall; 80% of participants had an ACB of zero and the maximum was 4.

**Table 1.**
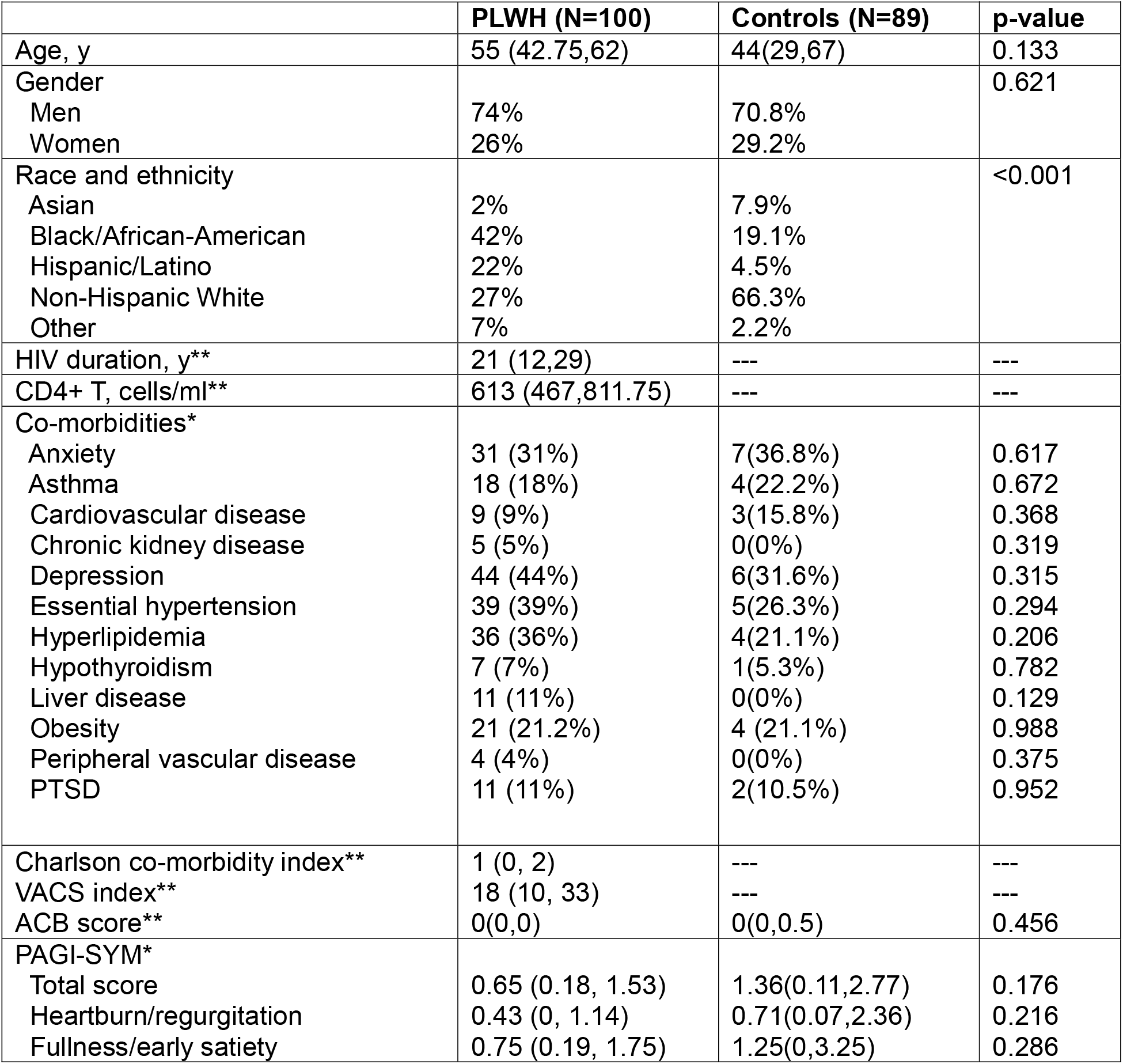
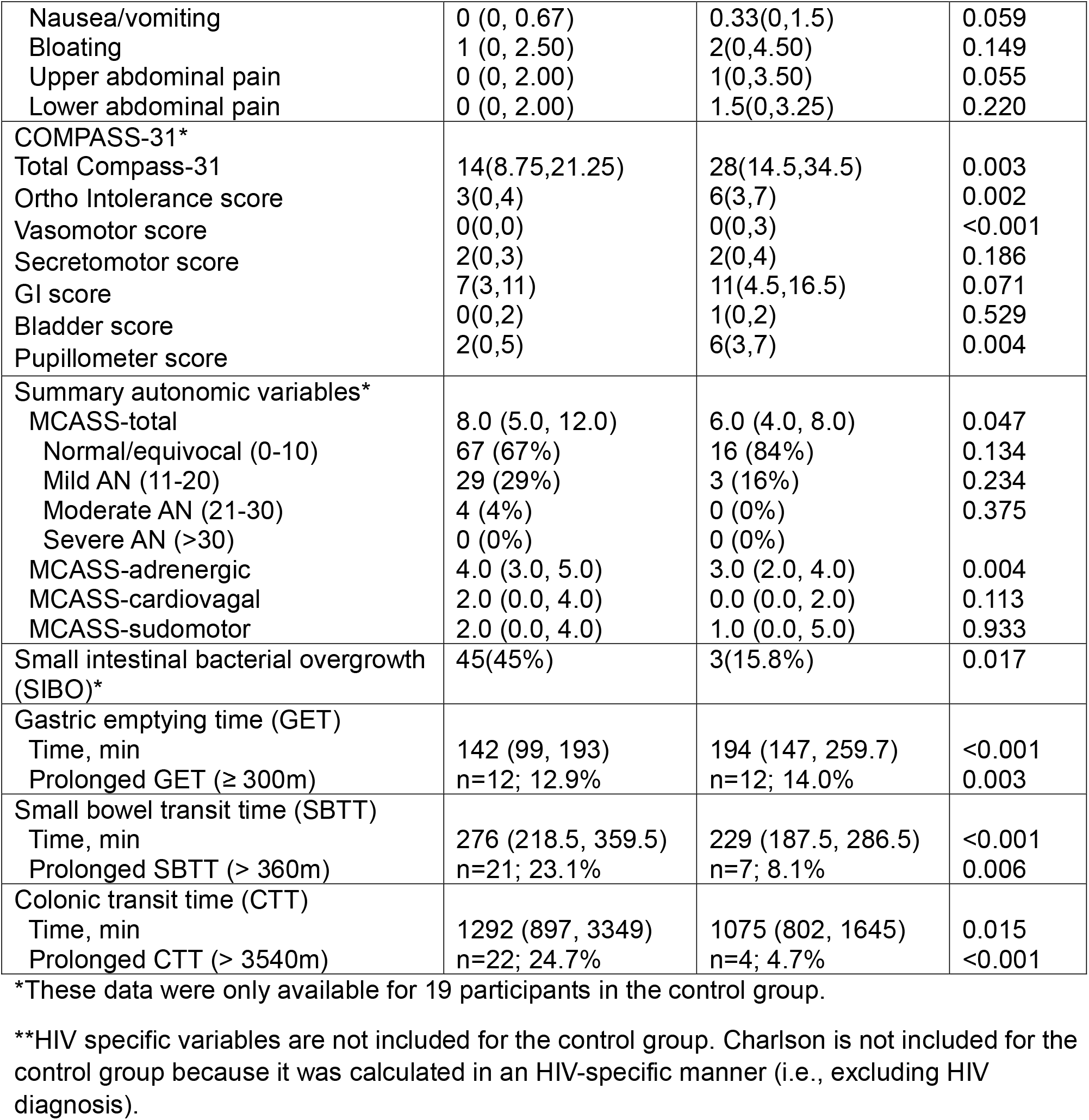
Participant demographics and GI functional parameters.

We were able to recruit 19 HIV-negative controls prior to the expiration date of our WMCs. As shown in Table 1, these prospective controls (which were the only controls who had autonomic and breath testing) had significantly better autonomic function than the PWH as evidenced by a lower MCASS score (p=0.047) and lower prevalence of SIBO (p=0.017). The age and gender composition of the control group overall (inclusive of prospective and historical controls) was similar to the group of PWH, however, the racial/ethnic composition was significantly different with more white participants among the controls.

### GI transit times (GET, SBTT, and CTT) differ between PWH and controls, and are correlated with patient-reported symptoms

Median GET was significantly shorter (p < 0.001), median SBTT was significantly longer (p < 0.001), and median CTT was significantly longer (p=0.015) in PWH compared to controls. This difference persisted in linear regression models with log-transformed GET (estimated coefficients=-0.366, p=0.002), log-transformed SBTT (estimated coefficients= 0.238, p=0.002) and log-transformed CTT as the outcome variables (estimated coefficients= 0.453, p=0.003). Regarding symptoms, among PWH, longer GET correlated with the total PAGI-SYM score r(91)=0.2, p=0.052, and with the fullness/early satiety sub-score r(91)=0.25, p=0.015. SBTT and CTT did not correlate with the PAGI-SYM.

### SBTT is associated with autonomic function in PWH; GET and CTT are not

The autonomic summary score, MCASS, was inversely correlated with SBTT, indicating that, unexpectedly, greater autonomic dysfunction overall was associated with shorter SBTT, r(89)=-0.23, p=0.026. The association persisted in multivariate linear regression adjusting for medications and medical co-morbidities (ACB score and VACS) with log-transformed SBTT as the outcome (estimated coefficients= -0.027, p= 0.031). The association was largely driven by the MCASS sudomotor sub-score (reflective of peripheral, non-cardiovascular, sympathetic function) which was also inversely associated with SBTT, r(89)=-0.24, p=0.023. SBTT was not correlated with the other two MCASS sub-scores: MCASS-adrenergic (reflective of cardiovascular sympathetic function) and MCASS-cardiovagal (reflective of parasympathetic/vagal function). GET and CTT were not correlated with MCASS or any of its sub-scores.

Regarding symptoms of autonomic and GI dysfunction there was a fairly strong correlation between the COMPASS-31 and PAGI-SYM total scores, r(98)=0.56, p<0.001. Multiple COMPASS-31 sub-scores were correlated with the PAGI-SYM total score. This included, not surprisingly, the COMPASS-31 GI sub-score, r(98)=0.47, p<0.001, but also three sub-scores that are calculated from non-GI symptoms and are typically associated with sympathetic dysfunction: orthostatic intolerance (e.g., dizziness on standing), r(98)=0.41, p<0.001; secretomotor (e.g., decreased sweat, tears and/or saliva), r(98)=0.36, p<0.001; gi score r(98)=0.47,p<0.001; and pupillomotor (e.g., light sensitivity) r(98)=0.33, p<0.001.

### Prolonged SBTT is associated with SIBO in PWH

Among the 21 participants with prolonged SBTT, 14 (67%) met diagnostic criteria for SIBO, compared to 29 of the 70 (41%) participants with normal SBTT, p = 0.042.

## Discussion

We found that GET was shorter and SBTT and CTT longer in PWH compared to healthy controls. SBTT was also inversely associated with MCASS, an overall indicator of HIV-AN; this association was driven by the MCASS sudomotor sub-score, which is a marker of peripheral sympathetic function. Thus, lower peripheral sympathetic function (as indicated by higher MCASS sudomotor sub-score) was associated with shorter SBTT, whereas parasympathetic/vagal function had no discernable relationship with SBTT. PWH with prolonged SBTT (>6h) were more likely to have SIBO than PWH with normal SBTT. Finally, although we were significantly underpowered to make comparisons between PWH and controls in anything but the WMC-based motility outcomes, it was notable that, both AN and SIBO were more common among PWH.

This is the first study of GI motility among PWH in the CART-era and the first to use the WMC in PWH. The WMC is a validated and FDA-cleared device that has been commonly used in clinical practice, and in research in a variety of disease states including cirrhosis, irritable bowel syndrome, post-operative ileus and diabetes.^33-36^ In people with diabetes and established sensorimotor polyneuropathy, the WMC demonstrated prolonged GET, SBTT and CTT compared to healthy controls. Presumably this was mediated by autonomic neuropathy (which commonly co-exists with sensorimotor polyneuropathy), although the authors did not specifically test autonomic function.^36^ In fact, ours is the first study to simultaneously measure GI motility using the WMC and autonomic function using a standardized laboratory-based battery.

Our first observation was that PWH had shorter GETs than controls. Although slowed gastric emptying has traditionally received more attention, rapid gastric emptying is increasingly recognized. Rapid gastric emptying is the most common GI motility abnormality in postural orthostatic tachycardia syndrome (POTS), another common autonomic disorder,^37^ and also occurs in diabetes.^38^ Gastric emptying rate is under complex neurohormonal control, designed to tightly regulate the rate of delivery of calories into the small intestine. This includes both excitatory and inhibitory parasympathetic/vagal circuits and “braking” hormones (e.g., GLP-1 and leptin).^38^ Also, more specific to PWH, the HIV-protein Tat has been shown to increase proximal GI motility in animal models through both direct and neurally-mediated mechanisms.^39,40^ Given this complexity it is not entirely surprising that we did not find a correlation between GET and autonomic function; more nuanced and multimodal methodology is likely necessary.

Our second observation was longer SBTT and CTT among PWH, and an association between longer SBTT and higher peripheral sympathetic function (as measured by the MCASS sudomotor sub-score). These findings are generally consistent with the expectation that increased sympathetic activity to the small intestine would be expected to slow motility,^41^ but that large intestinal motility would be more independent of the extrinsic ANS. However, the findings should be interpreted with caution due to the potential for inter-organ system variability in sympathetic activity. During a stress response, sympathetic activity may increase generally throughout the whole body. However, under normal physiologic conditions sympathetic activity to different organ systems is regulated by distinct mechanisms which are incompletely understood even in animal models, and almost entirely unexplored in humans. Thus, we do not know for certain whether sympathetic activity in the skin reflects sympathetic activity in the gut. Development and validation of practical techniques to assess regional sympathetic activity in humans is needed.

Our final finding, that PWH with prolonged SBTT were more likely to have SIBO, is consistent with prior (non-HIV-specific) literature.^14^ Future studies are needed to determine whether this corresponds to specific changes in the GI microbiome and ultimately to GI-derived antigenic stimulation which contributes to chronic inflammation in PWH.

This study has limitations. We were unable to prospectively recruit a large well-characterized control group because of time limitation due to the discontinuation of the WMC. The only data we had available for our historic controls were the WMC data themselves, age, sex and race/ethnicity. Thus, it is possible that there are other, unmeasured variables that contribute to the difference in GI transit times between the groups. PWH were recruited primarily from a single center in New York City and most had longer term HIV disease duration, so our results may not be representative of other populations of PWH.

In summary, this study demonstrates that altered GI motility occurs commonly in PWH and is associated with alterations in autonomic function and SIBO. Future work is warranted to more precisely characterize the autonomic pathophysiology and other clinical factors underlying altered GI motility in PWH and to understand how it might contribute to other well-established mechanisms by which the gut contributes to the overall health and well-being of PWH.

## Data Availability

All data produced in the present study are available upon reasonable request to the authors

## References

1. Brenchley JM, Price DA, Schacker TW, et al. Microbial translocation is a cause of systemic immune activation in chronic HIV infection. NatMed. 2006/12// 2006;12(12):1365–1371.

2. Pandrea I, Brooks K, Desai RP, Tare M, Brenchley JM, Apetrei C. I’ve looked at gut from both sides now: Gastrointestinal tract involvement in the pathogenesis of SARS-CoV-2 and HIV/SIV infections. Front Immunol. 2022;13:899559. doi:10.3389/fimmu.2022.899559

3. Tuddenham SA, Koay WLA, Zhao N, et al. The Impact of Human Immunodeficiency Virus Infection on Gut Microbiota α-Diversity: An Individual-level Meta-analysis. Clinical Infectious Diseases. 2019;70(4):615–627. doi:10.1093/cid/ciz258

4. Vujkovic-Cvijin I, Sortino O, Verheij E, et al. HIV-associated gut dysbiosis is independent of sexual practice and correlates with noncommunicable diseases. Nature Communications. 2020/05/15 2020;11(1):2448. doi:10.1038/s41467-020-16222-8

5. Sim JH, Mukerji SS, Russo SC, Lo J. Gastrointestinal Dysfunction and HIV Comorbidities. Curr HIV/AIDS Rep. Feb 2021;18(1):57–62. doi:10.1007/s11904-020-00537-8

6. Neild PJ, Nijran KS, Yazaki E, et al. Delayed gastric emptying in human immunodeficiency virus infection: correlation with symptoms, autonomic function, and intestinal motility. DigDisSci. 2000/08// 2000;45(8):1491–1499.

7. Sharpstone D, Neild P, Crane R, et al. Small intestinal transit, absorption, and permeability in patients with AIDS with and without diarrhoea. Gut. 1999/07// 1999;45(1):70–76.

8. Konturek JW, Fischer H, van der Voort IR, Domschke W. Disturbed gastric motor activity in patients with human immunodeficiency virus infection. ScandJGastroenterol. 1997/03// 1997;32(3):221–225.

9. Barnert J, Dumitrascu DL, Wienbeck M. Dyspepsia in AIDS is correlated to ultrasonographic changes of antral distension. EurJUltrasound. 2000/06// 2000;11(3):189–197.

10. Neild PJ, Evans DF, Castillo FD, et al. Effect of octreotide on small intestinal motility in HIV-infected patients with chronic refractory diarrhea. Dig Dis Sci. Dec 2001;46(12):2636–42. doi:10.1023/a:1012706908623

11. Zalar AE, Olmos MA, Piskorz EL, Magnanini FL. Esophageal motility disorders in HIV patients. Dig Dis Sci. May 2003;48(5):962–7. doi:10.1023/a:1023063916026

12. Robinson-Papp J, Sharma S, Simpson DM, Morgello S. Autonomic dysfunction is common in HIV and associated with distal symmetric polyneuropathy. JNeurovirol. 2013/04// 2013;19(2):172–180. doi:10.1007/s13365-013-0160-3

13. Camilleri M. Gastrointestinal motility disorders in neurologic disease. JClinInvest. 02/15/ 2021;131(4)doi:10.1172/JCI143771

14. Roland BC, Ciarleglio MM, Clarke JO, et al. Small Intestinal Transit Time Is Delayed in Small Intestinal Bacterial Overgrowth. J Clin Gastroenterol. 2015/08// 2015;49(7):571–576. doi:10.1097/MCG.0000000000000257

15. Robinson-papp J, Nmashie A, Pedowitz E, et al. Vagal dysfunction and small intestinal bacterial overgrowth: novel pathways to chronic inflammation in HIV. Aids. 2018/06/01/ 2018;32(9):1147–1156. doi:10.1097/QAD.0000000000001802

16. Robinson-Papp J, Nmashie A, Pedowitz E, et al. The effect of pyridostigmine on small intestinal bacterial overgrowth (SIBO) and plasma inflammatory biomarkers in HIV-associated autonomic neuropathies. J Neurovirol. Aug 2019;25(4):551–559. doi:10.1007/s13365-019-00756-9

17. Galligan JJ. HIV, opiates, and enteric neuron dysfunction. Neurogastroenterol Motil. Apr 2015;27(4):449–54. doi:10.1111/nmo.12539

18. Spencer NJ, Hu H. Enteric nervous system: sensory transduction, neural circuits and gastrointestinal motility. Nat Rev Gastroenterol Hepatol. Jun 2020;17(6):338–351. doi:10.1038/s41575-020-0271-2

19. Rao SS, Kuo B, McCallum RW, et al. Investigation of colonic and whole-gut transit with wireless motility capsule and radiopaque markers in constipation. Clin Gastroenterol Hepatol. May 2009;7(5):537–44. doi:10.1016/j.cgh.2009.01.017

20. Rao SS, Coss-Adame E, Valestin J, Mysore K. Evaluation of constipation in older adults: radioopaque markers (ROMs) versus wireless motility capsule (WMC). Arch Gerontol Geriatr. Sep-Oct 2012;55(2):289–94. doi:10.1016/j.archger.2012.04.003

21. Kuo B, McCallum RW, Koch KL, et al. Comparison of gastric emptying of a nondigestible capsule to a radio-labelled meal in healthy and gastroparetic subjects. Aliment Pharmacol Ther. Jan 15 2008;27(2):186–96. doi:10.1111/j.1365-2036.2007.03564.x

22. Camilleri M, Thorne NK, Ringel Y, et al. Wireless pH-motility capsule for colonic transit: prospective comparison with radiopaque markers in chronic constipation. NeurogastroenterolMotil. 2010/08// 2010;22(8):874-82, e233. doi:10.1111/j.1365-2982.2010.01517.x

23. Low PA. Composite autonomic scoring scale for laboratory quantification of generalized autonomic failure. Mayo ClinProc. 1993/08// 1993;68(8):748–752.

24. Robinson-Papp J, Sharma S, Dhadwal N, Simpson DM, Morgello S. Optimizing measures of HIV-associated neuropathy. Muscle & nerve. 2015/01// 2015;51(1):56–64. doi:10.1002/mus.24282

25. Rezaie A, Buresi M, Lembo A, et al. Hydrogen and Methane-Based Breath Testing in Gastrointestinal Disorders: The North American Consensus. Am J Gastroenterol. May 2017;112(5):775–784. doi:10.1038/ajg.2017.46

26. Robinson-Papp J, Sharma SK. Autonomic neuropathy in HIV is unrecognized and associated with medical morbidity. AIDS Patient Care STDS. 2013/10// 2013;27(10):539–543. doi:10.1089/apc.2013.0188

27. McGinnis KA, Justice AC, Moore RD, et al. Discrimination and Calibration of the Veterans Aging Cohort Study Index 2.0 for Predicting Mortality Among People With Human Immunodeficiency Virus in North America Clinical Infectious Diseases. 2021;75(2):297–304. doi:10.1093/cid/ciab883

28. Paudel M, Prajapati G, Buysman EK, et al. Comorbidity and comedication burden among people living with HIV in the United States. Curr Med Res Opin. Aug 2022;38(8):1443–1450. doi:10.1080/03007995.2022.2088714

29. Ramirez HC, Monroe AK, Byrne M, O’Connor LF. Examining the Association Between a Modified Quan-Charlson Comorbidity Index and HIV Viral Suppression: A Cross-Sectional Analysis of DC Cohort Participants. AIDS ResHumRetroviruses. 2023/12/01 2023;39(12):662–670. doi:10.1089/aid.2022.0186

30. Boustani M, Campbell N, Munger S, Maidment I, Fox C. Impact of Anticholinergics on the Aging Brain: A Review and Practical Application. Aging Health. 2008/06/01 2008;4(3):311–320. doi:10.2217/1745509X.4.3.311

31. Rentz AM, Kahrilas P, Stanghellini V, et al. Development and psychometric evaluation of the patient assessment of upper gastrointestinal symptom severity index (PAGI-SYM) in patients with upper gastrointestinal disorders. Qual Life Res. 2004/12// 2004;13(10):1737–1749. doi:10.1007/s11136-004-9567-x

32. Sletten DM, Suarez GA, Low PA, Mandrekar J, Singer W. COMPASS 31: a refined and abbreviated Composite Autonomic Symptom Score. Mayo Clin Proc. 2012/12// 2012;87(12):1196–1201. doi:10.1016/j.mayocp.2012.10.013

33. Chander Roland B, Garcia-Tsao G, Ciarleglio MM, Deng Y, Sheth A. Decompensated cirrhotics have slower intestinal transit times as compared with compensated cirrhotics and healthy controls. J Clin Gastroenterol. Nov-Dec 2013;47(10):888–93. doi:10.1097/MCG.0b013e31829006bb

34. DuPont AW, Jiang ZD, Harold SA, et al. Motility abnormalities in irritable bowel syndrome. Digestion. 2014;89(2):119–23. doi:10.1159/000356314

35. Vilz TO, Pantelis D, Lingohr P, et al. SmartPill® as an objective parameter for determination of severity and duration of postoperative ileus: study protocol of a prospective, two-arm, open-label trial (the PIDuSA study). BMJ Open. Jul 8 2016;6(7):e011014. doi:10.1136/bmjopen-2015-011014

36. Farmer AD, Pedersen AG, Brock B, et al. Type 1 diabetic patients with peripheral neuropathy have pan-enteric prolongation of gastrointestinal transit times and an altered caecal pH profile. Diabetologia. Apr 2017;60(4):709–718. doi:10.1007/s00125-016-4199-6

37. Loavenbruck A, Iturrino J, Singer W, et al. Disturbances of gastrointestinal transit and autonomic functions in postural orthostatic tachycardia syndrome. Neurogastroenterol Motil. Jan 2015;27(1):92–8. doi:10.1111/nmo.12480

38. Goyal RK, Cristofaro V, Sullivan MP. Rapid gastric emptying in diabetes mellitus: Pathophysiology and clinical importance. J Diabetes Complications. Nov 2019;33(11):107414. doi:10.1016/j.jdiacomp.2019.107414

39. Ngwainmbi J, De DD, Smith TH, et al. Effects of HIV-1 Tat on enteric neuropathogenesis. J Neurosci. 2014/10/22/ 2014;34(43):14243–14251. doi:10.1523/JNEUROSCI.2283-14.2014

40. Esposito G, Capoccia E, Gigli S, et al. HIV-1 Tat-induced diarrhea evokes an enteric glia-dependent neuroinflammatory response in the central nervous system. Sci Rep. Aug 10 2017;7(1):7735. doi:10.1038/s41598-017-05245-9

41. Camilleri M. Gastrointestinal motility disorders in neurologic disease. J Clin Invest. Feb 15 2021;131(4)doi:10.1172/JCI143771

